# Optimizing Wastewater Surveillance Sites for COVID-19 Hospitalization Forecasting Across U.S. States

**DOI:** 10.64898/2026.07.20.26358167

**Authors:** Gursharn Kaur, Jiangzhuo Chen, Abhijin Adiga, Aniruddha Adiga, Baltazar Espinoza, Bryan Lewis, Madhav Marathe, Srinivasan Venkatramanan, Andrew Warren, Anil Vullikanti

## Abstract

In this study, we analyze viral load data from wastewater treatment plants (WWTPs) across multiple U.S. states and address the challenge of selecting an optimal subset of sites to improve COVID-19 hospitalization forecasts. Using forward (greedy) regression with a baseline ARIMA model, we identify the most informative WWTPs that enhance forecast accuracy while reducing the number of sampling locations. Our analysis, based on NWSS data, shows that the optimal number of sites typically ranges from 2-8, though some states, including NY, IL, and WI, benefit from a larger set (10-20). We also leverage a Virginia-level digital twin model specifically designed for wastewater data modeling and analysis and our results show that for different parameter settings, that forecast accuracy can be achieved with strategically chosen small number of sites, providing scalable and cost efficient wastewater surveillance.

## 1 INTRODUCTION

Wastewater-based epidemiology (WBE), which involves monitoring pathogen in waste water, has become an essential component of national infectious disease surveillance across the world for many diseases, including SARS-CoV-2, influenza and monkeypox, e.g., (Amman et al. 2022; Chen et al.; Lopez et al. 2024). In the U.S., the National Wastewater Surveillance System (NWSS) was set up to provide measurements of SARS-CoV-2 RNA concentrations and other pathogen across hundreds of wastewater treatment plants (WWTPs) nationwide. These measurements capture viral shedding from both symptomatic and asymptomatic individuals and often precede changes in clinical indicators such as hospital admissions. Consequently, wastewater signals offer a valuable, unbiased, and community-wide early-warning system for public health decision-making. Wastewater surveillance has also been found useful for monitoring other kinds of materials, such as pharmaceuticals and illicit drugs, e.g., (Bischel et al. 2015).

Although wastewater surveillance is generally less expensive than other forms of disease surveillance, and many sites were set up during COVID-19, implementation and sustained operation still incur substantial costs. Costs of surveillance include initial infrastructure setup and for regular operation, which include logistical and human resource costs for sampling and sequencing on a regular basis for an extended period of time. As a result, there has been a lot of interest in choosing wastewater surveillance sites in an optimal manner, e.g., (Wang et al. 2024; Calle et al. 2021). While most of the prior work emphasize maximizing population coverage by the chosen subset; we propose a complementary approach that focuses on identifying a minimal subset of sites that improve hospitalization forecasting.

This work was also motivated by discussions with colleagues at the U.S. Centers for Disease Control and Prevention (CDC), who highlighted the practical challenges of sustaining large-scale wastewater surveillance networks. In particular, understanding how many-and which-sites are truly necessary for effective public-health decision-making emerged as a key question for ongoing surveillance planning. These conversations reinforced the need for data-driven approaches to site selection that improve forecast performance with operational feasibility.

In this paper, we study the Optimal Wastewater Site Selection problem (OptWWSiteSelect), which aims to identify an optimal subset of wastewater monitoring sites such that epidemic forecasting performance is maximized using data collected from the selected sites. A formal mathematical definition of the problem is provided in Section 2. We use observed wastewater viral load data from NWSS for all states. Despite the expanded geographic coverage of NWSS, forecasting COVID-19 hospitalizations directly from wastewater remains challenging. Substantial heterogeneity exists across states in terms of sewershed scale, WWTP sampling frequency, laboratory quantification protocols, population mobility, and hospitalization reporting practices. These factors induce variability in both the strength and the lead time of the wastewater–hospitalization relationship. Moreover, the predictive value of wastewater may diminish with longer forecast horizons, raising the need for systematic evaluation of wastewater-driven forecasting under different lead times.

OptWWSiteSelect can be solved by retrospective analysis; however, a significant limitation is that the chosen sites might not be effective for new kinds of epidemic scenarios, e.g., importation in new regions, and infections within a new subpopulation, which did not occur in retrospective data. Such scenarios arose multiple times during the COVID-19 pandemic, where new variants emerged in different subpopulations, and changed the disease dynamics. Since the initial setup cost in wastewater surveillance is very high, it is important to find solutions that would be effective not just for retrospective viral loads, but new epidemic scenarios. Further, the optimal solution for one scenario might not be optimal for a different scenario.

In this work, we address this problem in two ways. First, we formalize OptWWSiteSelect using the notion of robust optimization, which involves finding a solution whose performance is “good” over all scenarios. Second, we use a novel *Wastewater Digital Twin (WDT)* for generating a broad class of simulated scenarios. WDT models individual level disease states within a population, along with a wastewater network, and viral loads at each site based on disease states within the population. It also models different kinds of scenarios, such as importations in different regions, as well as interventions such as vaccination. Our results are summarized below.

### 1. Optimal site selection with NWSS data

We develop a nationwide forecasting framework that integrates NWSS wastewater measurements with auto-regressive time-series components and a greedy strategy for OptWWSiteSelect. We use this to quantify the predictive contribution of wastewater data to short-term hospitalization forecasts across all U.S. states reporting both WWTP-level NWSS data and state-level hospitalization counts. For each state, we construct forecasts for four prediction horizons: *h* ∈ {1, 2, 3, 4} corresponding to 1-to 4-week-ahead prediction of weekly COVID-19 hospital admissions. Each forecast horizon is treated as an independent *scenario*. This formulation enables a unified comparison of wastewater predictive performance across different lead times. We found that for several major states, the optimal number of sites for improving the prediction accuracy is quite small, typically between 2 and 8. In states such as New York, Illinois, and Wisconsin, a higher number of sites was required to achieve better accuracy across four-week hospitalization forecasts.

Further, the observed underlying distribution of population served by individual WWTPs within states is heavily skewed toward smaller proportions, with only 21 sites across all states serving more than 30% of a total state’s population served. In contrast to coverage-focused approaches (e.g., (Wang et al. 2024; Calle et al. 2021)), our retrospective analyses shows that the optimal site selection typically include sites with up to 20% coverage. This highlights the importance of careful site selection rather than purely relying on facility size.

### 2. Robust solutions using Wastewater Digital Twin Simulations

To further evaluate the robustness and generalizability of the site-selection strategy, we study our solution using a wastewater digital twin (WDT) developed for the state of Virginia. WDT provides a rich ensemble of simulated scenarios for Virginia, which models viral loads at all wastewater sites and hospitalization rates at the state level (Section 6). We use the WDT to generate a class of scenarios, and then apply the same modeling approach and site-selection pipeline as in the NWSS analysis to evaluate 1–4 week ahead forecasts. Importantly, the digital twin allows us to assess the stability of selected sites under varying parameter settings. We observe that the state level Hospitalization counts and wastewater viral load (WVAL) data were closely aligned with correlation between hospitalizations and WVAL from multiple WWTPs were significant, mostly ranging between 0.3 and 0.8. We observe that even under three different scenarios (and multiple replicates), the number of optimal sites remain very small compared to the total number of sites sampling.

Further, we evaluate performance using a rolling-window forecasting setup to mimic real-time deployment. We compare the proposed site selection approach with LASSO-based approaches to understand trade-offs between forecast accuracy and model complexity. Models were trained using an expanding window with a minimum training length of 30, and forecasts were generated for horizons up to four weeks ahead. The greedy ARIMA-X model typically includes no more than 20% of all available sites in order to improve the predictive performance. In contrast, LASSO based methods tend to select a much larger subset of sites –often upto 80%– while achieving comparable forecasting performance.

## 2 PROBLEM SETUP: OPTWWSITESELECT FOR WASTEWATER SURVEILLANCE

Let *V* denote the set of wastewater sites in a given state. Let ℛ denote a set of scenarios, e.g., different simulated outbreak conditions and a forecast horizon. For each scenario *r* ∈ ℛ, let *X*_*j*_(*t*) denote the time series of *viral activity levels* (VALs) from a wastewater site *j* ∈ *V* and *Y* ^*r*^ (*t*) denote the corresponding hospitalization time series. For each scenario *r*, we first consider a baseline (ARIMA) forecast 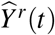 of *Y* ^*r*^(*t*), which is based solely on past hospitalization data. We define the forecast residuals as

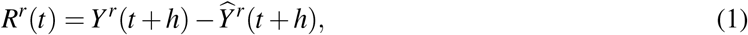

where 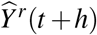 denotes the scenario-specific baseline forecast and *h* is the forecast horizon associated with scenario *r*. To assess the additional predictive value of wastewater data, we model these residuals using VAL signals. For a subset of sites *S* ⊆ {1,…, *P*} we consider the linear regression model

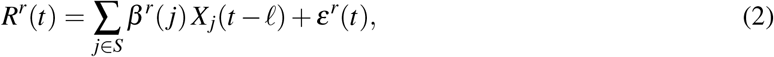

where *ℓ* is a fixed lag, *β*^*r*^(*j*) are scenario-specific regression coefficients. We evaluate a predictor set *S* using the adjusted coefficient of determination (Adj *R*^2^), which is defined as

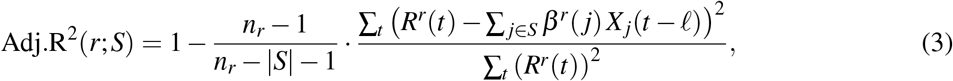

where *n*_*r*_ denote the number of observations available under scenario *r*.

### Optimal Wastewater Site Selection Problem (OptWWSiteSelect)

For a fixed scenario *r*, the site selection problem involves finding a subset of sites that maximizes Adj.R^2^. That is

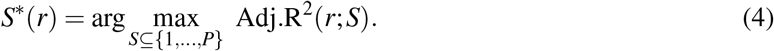

### Robust Wastewater Site Selection

The robust objective is to select a subset of sites that performs well across all scenarios. That is

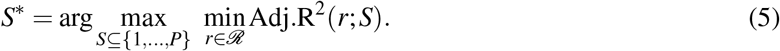

## 3 RELATED WORK

Wastewater surveillance is a powerful tool that can help detect outbreaks early and guide public health actions (Larson et al. 2020; Nourinejad et al. 2021). Compared to testing individuals, it can save costs and focus efforts where they’re most needed. Several studies have formulated site selection as sensor placement over network structure, to maximize the detection probabilities of contamination or viral presence (Mac Mahon et al. 2022). Related research in water distribution systems similarly formulates optimal sensor placement as a multi-objective optimization problem, with common objectives including maximizing coverage, minimizing detection time, reducing the number of sensors, and limiting population exposure (Berry et al. 2006; Krause et al. 2008).

Recent studies by (Eugene et al. 2025) have shown that by carefully choosing which sites to monitor, it is possible to get reliable results without having to test everywhere, making surveillance both smarter and more efficient. While this study for Honk Kong explored reductions in sampling sites by selecting subsets based primarily on population coverage, by choosing sites in descending order of coverage, we adopt a prediction-driven approach on wastewater site selection. Specifically, we leverage viral load data from all WWTPs to predict hospitalizations and use forward regression to identify the most informative sites. Further, unlike the population-based selection, our method quantifies the trade-off between the number of sites and forecasting performance, providing a complete picture of which WWTPs contribute most to prediction accuracy while maintaining scalability across multiple U.S. states.

## 4 METHODS

We employ a multi-stage modeling framework that combines autoregressive integrated moving average (ARIMA) components with a greedy linear selection mechanism for wastewater predictors. The objective is to quantify the incremental predictive value of wastewater sites for short-term hospitalization forecasting, while explicitly controlling for model complexity.

### Baseline Time Series Model

Let *Y*(*t*) be the weekly hospitalization time series for a given scenario. To capture intrinsic temporal dependence in *Y*(*t*), we fit a univariate autoregressive integrated moving average (ARIMA) model, where model orders (*p, d, q*) are selected using Akaike Information Criterion (AIC) minimization. The fitted ARIMA model serves as a baseline that captures intrinsic temporal dynamics of hospital admissions, and the ARIMA forecast residuals (as in equation (1)) represent the variation not explained by autoregressive structure alone.

### Wastewater-Based Predictor Selection

To incorporate wastewater signals, we model the ARIMA residuals as a function of WWTP-level VALs using a linear regression framework as in equation (2). Given the empirically observed correlation between wastewater VALs and hospitalizations, linear regression provides a simpler and effective modeling approach. We apply a forward (greedy) variable selection approach to identify a subset of optimal wastewater predictors. Starting with an empty model. At each step, the WWTP variable that provides the greatest improvement in adjusted *R*^2^ model is selected. Statistical significance of incremental improvement is evaluated using an F-test and the procedure continues until no additional predictor yields a statistically significant improvement. Final forecasts are generated by augmenting the ARIMA baseline with a linear regression model fitted on the selected wastewater predictors. We refer to this modeling framework as **greedy ARIMA-X**.

Since the coefficient of determination (R^2^) automatically increases with additional predictors, we use adjusted R^2^, as measure of accuracy, which also accounts for model complexity. Consequently, maximizing Adj.R^2^ is equivalent to minimizing the residual sum of squares (RMSE) with an explicit penalty for each additional predictor, and thus it closely correspond to optimizing forecast error subject to variable selection cost. The use of Adj.R^2^ and greedy subset selection is further motivated by theoretical guarantees for subset selection under R^2^-based objectives, where greedy procedures admit bounded approximation performance (Das and Kempe 2018).

### Evaluations with Observed and Simulated Data

For observed surveillance data, we evaluate model fit using complete historical data to identify informative predictors. However, rolling-window evaluation is challenging in real-time settings because wastewater measurements are often incomplete or unavailable during portions of the evaluation period. To enable out-of-sample evaluation and a fair comparison of model complexity, we leverage data generated from a wastewater digital twin, which provides complete and temporally aligned measurements across all wastewater sites. Using these simulated data, we conduct rolling-window forecasting experiments and analyze the predictive performance and number of optimal sites with the greedy ARIMA-X framework. For comparison, we also analyze LASSO regression models that include lagged hospitalizations and VALs as predictors.

## 5 RESULTS: SITE SELECTION WITH OBSERVED DATA

We construct a weekly, state-specific dataset for all U.S. states that integrates wastewater surveillance signals with hospitalization outcomes. The resulting dataset is temporally aligned and serves as the basis for the analyses described below. The data components are as follows.

### Wastewater surveillance data

We use the publicly available NWSS dataset^1^ from all wastewater treatment plants (WWTPs) operating within state, along with associated county information and the population served by each facility. We used the population served by each WWTP to analyze our model outputs. For each week, we aggregate all observations from each WWTP by computing the mean, producing a single weekly value per variable. To standardize the viral load measurements, we convert the raw data into VALs following the NWSS normalization procedure. VALs are computed by log-transforming the viral load, then centering and scaling based on a baseline derived from the 10th percentile and its standard deviation over the preceding six months. For periods with less than six months of historical data, a moving 10th percentile and SD is used to calculate VALs.

### Hospitalization data

The weekly COVID-19 hospitalization data is also publicly available^2^. The time series begins in January 2020 and, for several states data is updated through April 2024. We consider weekly number of confirmed COVID-19 hospital admissions in a state as a target variable, denote by *Y* (*t*).

After aligning wastewater and hospitalization records to a common weekly calendar, we obtain a unified predictor–response dataset, where each observation at week is given by 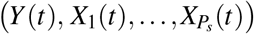. This dataset forms the basis for all subsequent state-level modeling and forecasting analyses. The number of wastewater treatment plants (WWTPs) varies substantially across states, ranging from 2 to 159, with a median of 20 sites per state. States such as New York (159), Illinois (72), and Michigan (60) have a large number of sites, while states like Virginia and California have 28 unique WWTPs. However, not all sites report data consistently, resulting in missing values. We address this by first applying linear interpolation between consecutive weeks and then excluding sites with more than 40% missing observations over the study period.

### Association Between Wastewater and Hospitalizations

Figure 1 shows the weekly hospitalization time series alongside wastewater viral load (log-scale) for selected states. We observe strong temporal alignment between the two signals, particularly during the 2023–2024 period. In several states, wastewater viral load acts as a leading indicator of hospitalization trends, suggesting its utility as an early signal for forecasting. To quantify the relationship between wastewater signals and hospitalizations, we compute the Pearson correlation between *Y*_*s*_(*t*) and viral load measurements from each WWTP. By considering the WWTPs per state with statistically significant correlations (*p* < 0.05) and magnitude greater than 0.2, we observe that a more than 70% fraction of WWTPs exhibit moderate to strong correlation with hospitalization time series, indicating that wastewater data may have predictive information. This also confirms our model choice, where we regress the residuals from a baseline ARIMA model on the WVAL time series from selected WWTPs to capture additional variation not explained by the baseline model.

**Figure 1.**
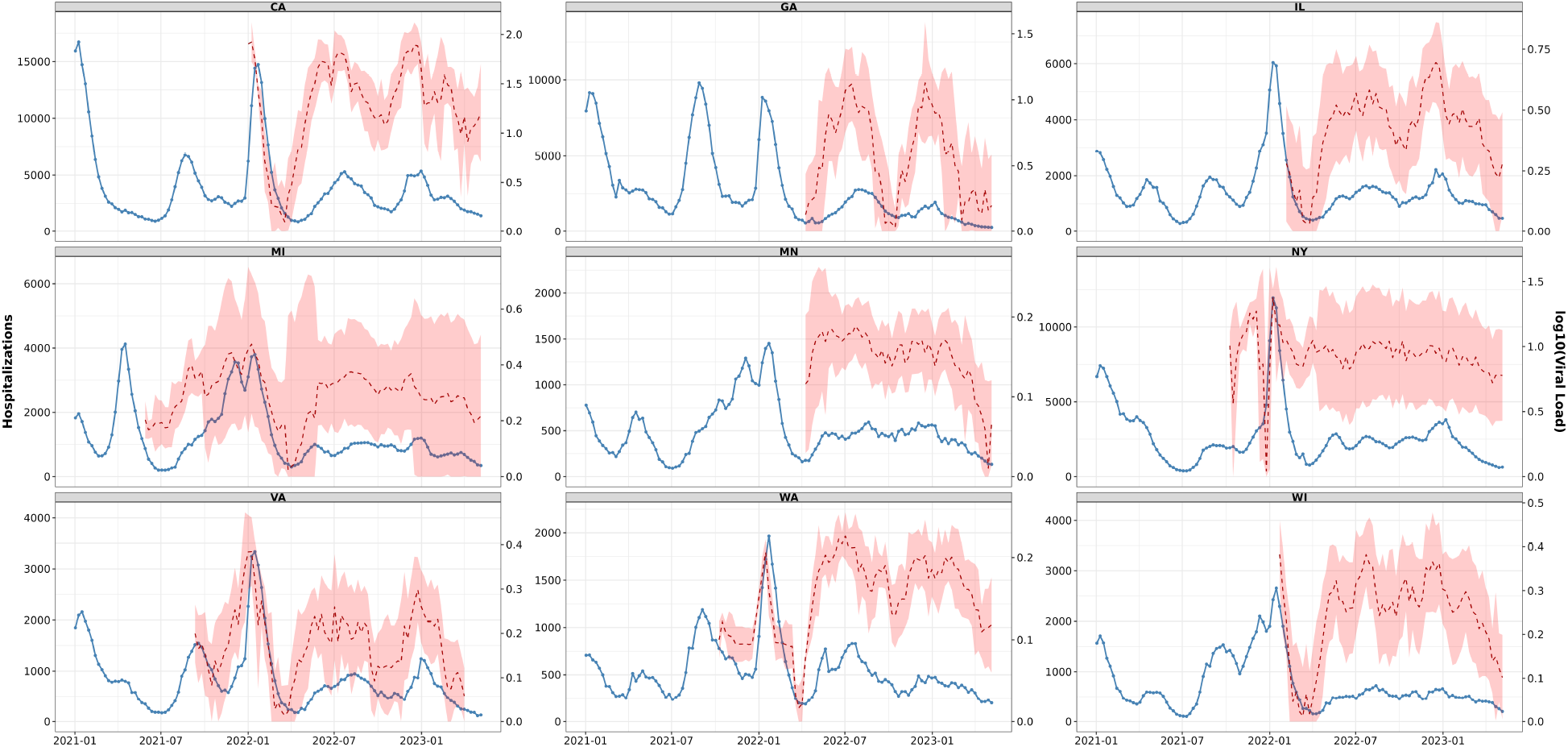
Weekly COVID-19 hospitalizations and wastewater viral load (log_10_ scale) across selected U.S. states. Wastewater viral load is shown as the mean across sites within each state, with shaded regions representing the 95% confidence interval.

#### 5.1 Selected Wastewater Surveillance Sites

We evaluate the proposed site selection approach across multiple forecast horizons. Figure 2 illustrates how model performance, measured by Adj.R^2^, changes with the number of selected WWTP predictors. The optimal number of WWTP sites, highlighted in boxes, for each forecast horizon, corresponds to the subset selected by the proposed forward selection algorithm. This shows that a relatively small number of carefully chosen sites can achieve strong predictive performance while avoiding overfitting.

**Figure 2.**
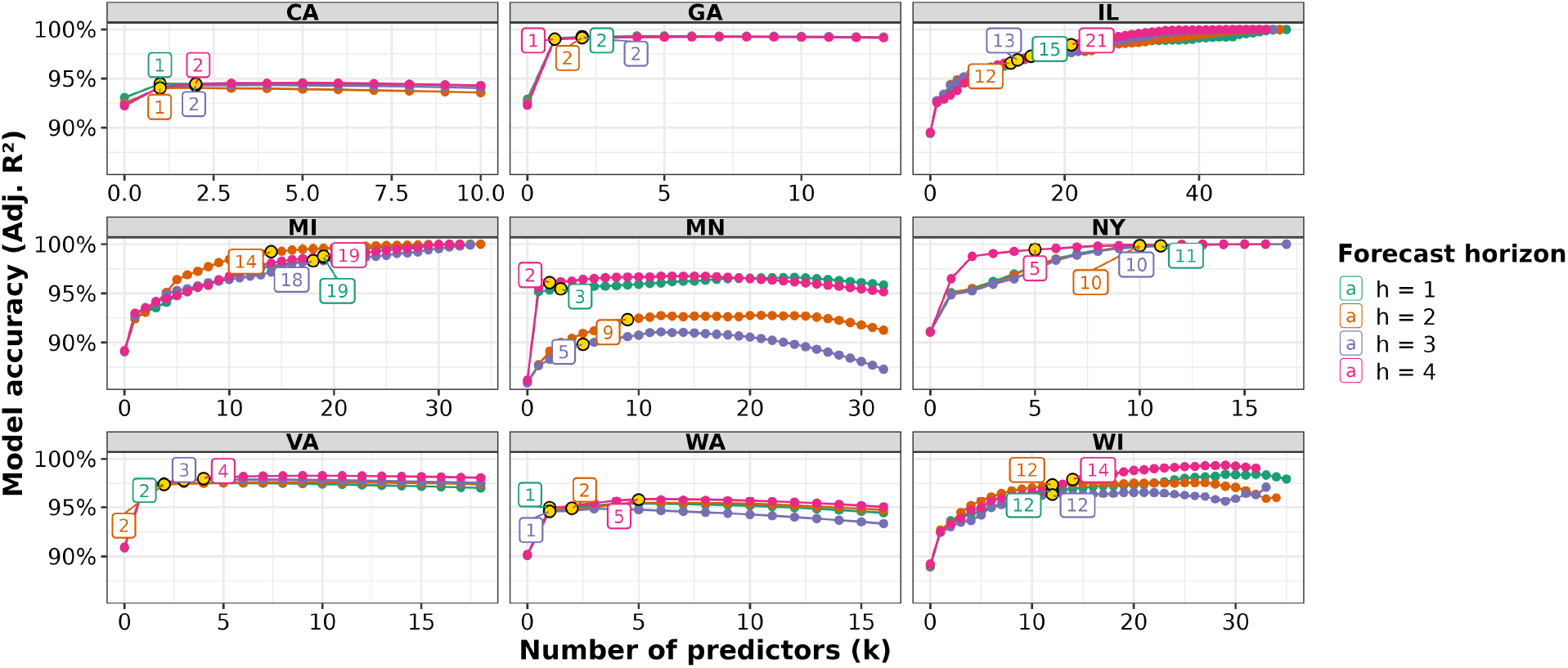
Model accuracy as a function of number of WWTPs included in the greedy ARIMA-X model. Each panel correspond to one of 9 selected states, and within each panel, lines represent forecast horizons *h* (1-4 weeks ahead). Model accuracy is measured with Adjusted R^2^ and the labels represent the optimal number of selected sites for the given horizon.

The distribution of the selected sites by forecast horizon across different states is shown in Figure 3, which summarizes the optimal number of wastewater treatment plants selected for each prediction horizon. Across all horizons and states, the number of selected sites is generally small, most often ranging between 2 and 8, indicating that only a limited subset of WWTPs is sufficient to add value to hospitalization forecasting in most states. We also assessed population coverage of the WWTPs selected by the forecasting models and found that most selected sites typically serve upto 20% population. While high coverage facilities are often included when present, optimal site selection is primarily driven by the sites that provide the most informative viral load signals for predicting hospitalizations rather than by facility size.

**Figure 3.**
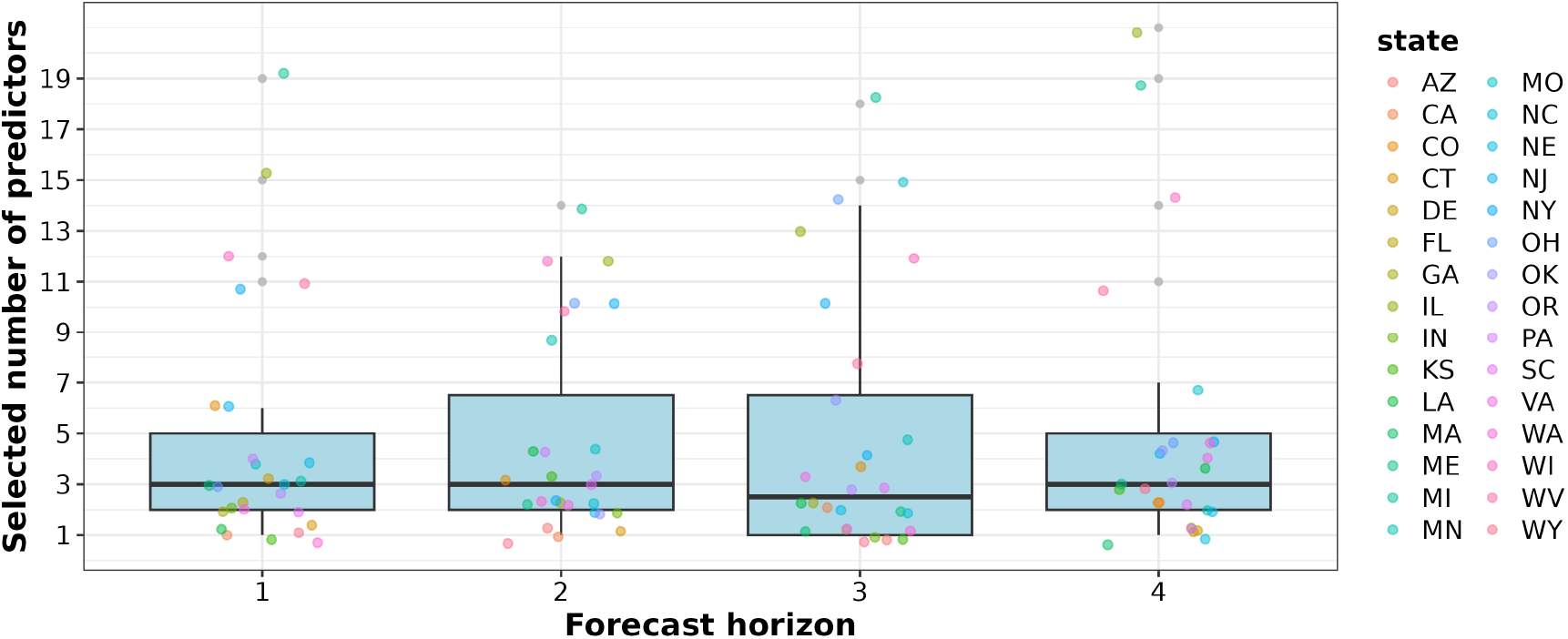
Distribution of the optimal number of selected wastewater treatment plants (|*S*^∗^|) across forecast horizons. For each horizon (1-4 weeks ahead), the boxplot summarizes number of sites selected across all states. Individual points correspond to state-specific optimal site counts.

## 6 VIRGINIA WASTEWATER DIGITAL TWIN

We generate Virginia synthetic wastewater data as an additional layer in the digital twin of Virginia, using an individual-level agent-based model, based on real NWSS WWTP location and capacity information from CDC.

### Digital twin

A Digital twin of a region captures the people in the region with individual-level demographic attributes, their partition into households with household attributes, an activity sequence for each individual, and a set of residence and activity locations where people conduct their activities. The location assignment of activities formulates a people-location network *G*_*PL*_, consisting of edges (*v, ℓ, a, t*_*b*_,*t*_*e*_), representing individual *v* visiting location *ℓ* from time *t*_*b*_ to time *t*_*e*_ for the purpose of activity type *a*. Each location *ℓ* is associated with latitude and longitude coordinates. The mapping of people’s activities to locations allows us to keep track of the location of any individual at any time, which is used to map their viral sheddings to physical locations. The construction of the digital twin is carried out so that the synthetic population closely resembles their real counterparts on dimensions relevant for epidemic scenarios. A more detailed overview of the construction methodology and their validation is provided in (Mortveit et al. 2020).

### Pandemic scenario

We first extend our Virginia digital twin with a synthetic pandemic scenario, defined as follows. The scenario consists of (*i*) a COVID-19 disease model with age stratification and waning immunity, (*ii*) initializations based on state level importations data, and (iii) vaccination intervention based on state level vaccine administration data from CDC. The disease model is illustrated in Figure 4. It features three infectious states: asymptomatic (*A*), presymptomatic (*P*), symptomatic (*I*); three susceptible states: naively susceptible (*S*), susceptible after waning (*W*), vaccinated (*V*); as well as a hospitalized state (*H*), a death state (*D*), and a recovered state (*R*). The model incorporates early transition states that represent the clinical progression of symptomatic individuals (*I*) toward different outcomes: hospitalization transition (*hM*), death transition (*dM*), and recovery transition (*rM*). Moreover, critical clinical stages for those on a terminal path are incorporated according to the specific track leading to death: hospitalized individuals (*dH*) and a terminal state for patients on mechanical ventilation (*dVent*). These compartments enable accurately map individual viral shedding to physical locations based on an individual’s stage of illness and activity. Any node that has just recovered from infection becomes susceptible again after *d*_*R*→*W*_ days, where *d*_*R*→*W*_ is sampled from an exponential distribution of mean 6 months. Vaccinated nodes are in V state with 95% protection against infection.

**Figure 4.**
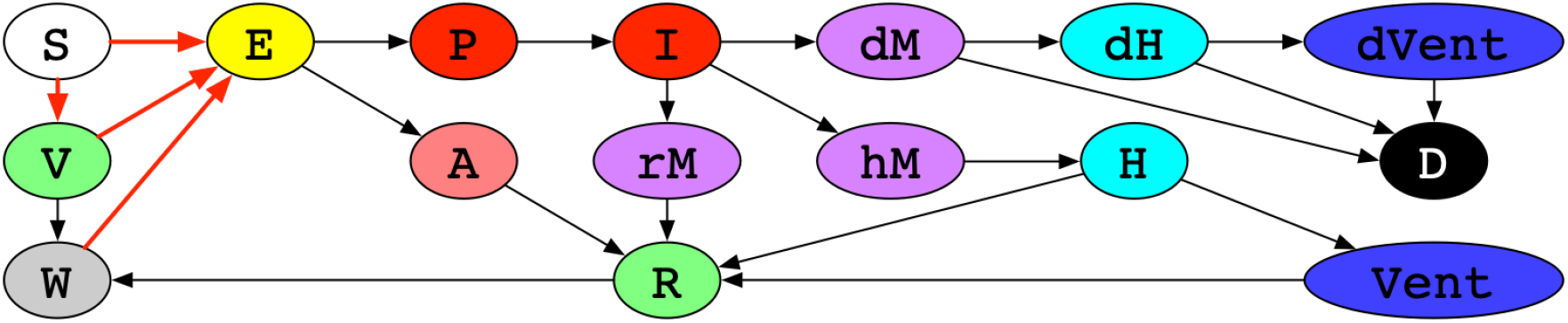
COVID-19 disease model with age stratification and waning immunity.

**Figure 5.**
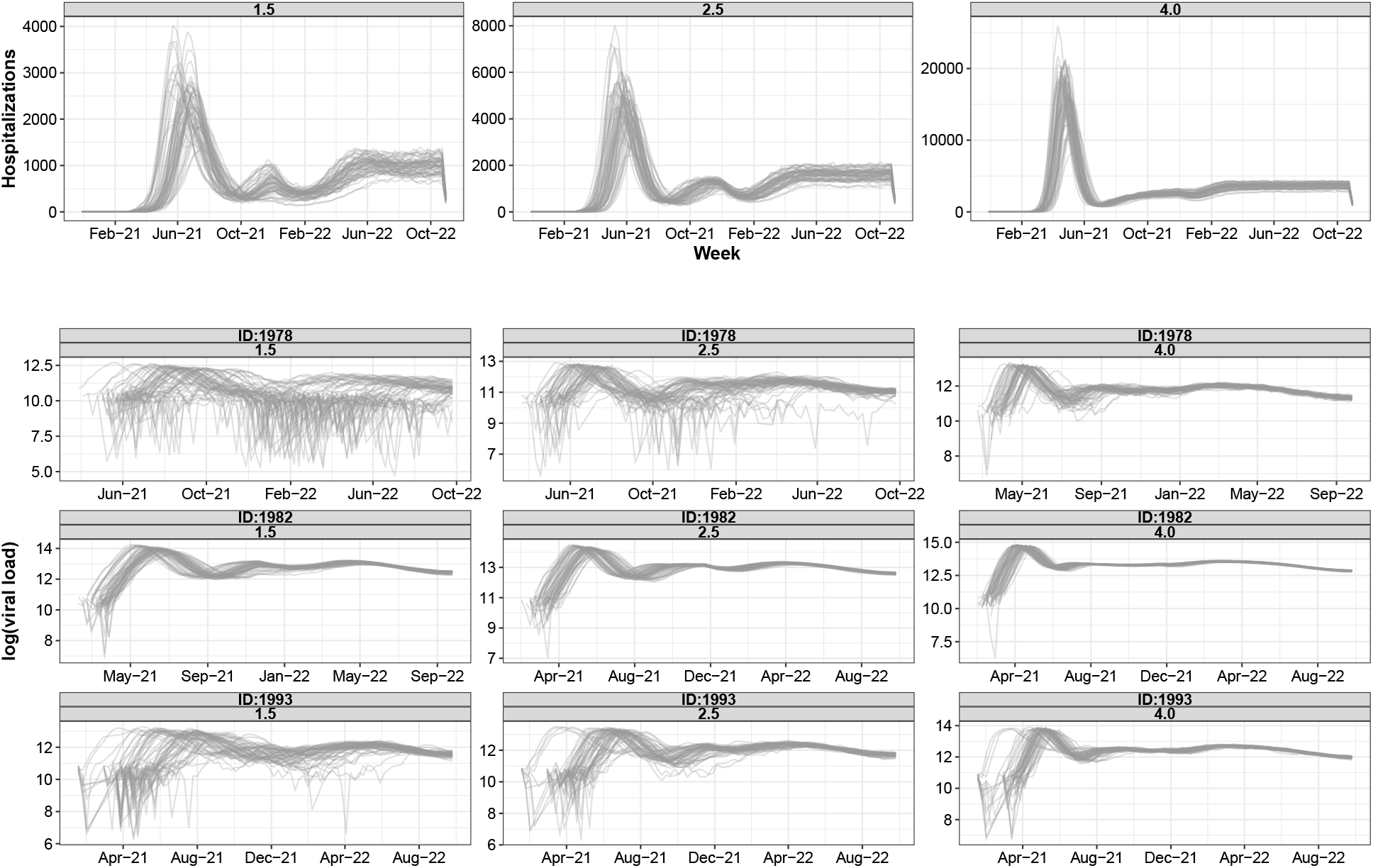
Simulation outputs from the wastewater (WW) digital twin model across 60 stochastic replicates. The top panel shows hospitalization trajectories for three different basic reproduction numbers (*R*_0_ = 1.5, 2.5, and 4), where each curve represents a single replicate. The bottom panel shows wastewater viral load for three selected sewersheds, with each panel displaying 60 replicate trajectories.

We calibrate transmissibility *τ* of in the disease model against different *R*_0_ values {1.5, 2.5, 4.0}. We initialize the agent-based model by randomly seed infections in the whole state population based on state level time series of importations. The scenario includes three cells, corresponding to different *R*_0_ values {1.5, 2.5, 4.0}, and lasts for 700 days. For each cell we run our agent-based model 60 replicates. Using our agent-based model EpiHiper (Chen et al. 2025), the scenario generates *synthetic pandemic data*, consisting of the health state of every individual on every day. We aggregate it to obtain state level time series of hospitalizations.

EpiHiper is a high performance computing-based modeling framework for epidemics ((Hoops et al. 2021; Bhattacharya et al. 2024; Moon et al. 2024; Chen et al. 2025)). It can simulate epidemic dynamics over large-scale networks while supporting modulation of the epidemic trajectory through user-programmable interventions, including pharmaceutical ones (such as vaccination) and non-pharmaceutical ones (such as school closure). Its software architecture is a hybrid MPI/OpenMP design implemented in C++ for high performance. The highly efficient and scalable implementation allows EpiHiper to routinely handle simulations on digital twins of national scale populations, such as the ditial twin of the USA population, which consist of hundreds of millions of people and their detailed contact networks. EpiHiper is designed to allow analyzing highly detailed what-if scenarios under a range of interventions and disease strains such as for COVID-19. It has been used to support policymakers and epidemiologists for planning and response efforts, such as the CDC Scenario Modeling Hub for COVID-19 response (Chen et al. 2024).

### Synthetic wastewater data

Using the Virginia digital twin with the synthetic pandemic extension, we can further extend to get synthetic wastewater data as follows: (*i*) We apply a temporal viral shedding model from (Phan et al. 2023) to generate the shedding quantity of each infected individual on each day. (*ii*) Based on the people-location network *G*_*PL*_, we determine the locations of shedding probabilistically. (*iii*) Based on the counties each WWTP serves, we randomly assign individuals to WWTPs, and route their viral shedding to their corresponding WWTPs with realistic delay and decay. (*iv*) Finally, we obtain aggregate viral load arriving at each WWTP on every day. Time series of state level hospitalizations and that of WWTP level viral activity levels (VAL) are used to test the robustness of our solutions to OptWWSiteSelect.

Across all 60 stochastic replicates of the wastewater digital twin model, the mean correlation between wastewater viral activity levels (VAL) from wastewater treatment plants (WWTPs) and hospitalizations ranged from −0.47 to 0.59, with a median of 0.34. The interquartile range across values of *R*_0_ was 0.17–0.44, indicating consistently positive correlations across WWTPs and parameter settings.

#### 6.1 Selected Wastewater Surveillance Sites with Digital Twin

As defined in Section 2, each scenario can correspond to the combination of epidemic parameters or forecast horizons. Here we analyze the robust solution, defined as the optimal subset of sites that minimizes forecast error jointly across all forecast horizons. Figure 6 illustrates, how frequently a given number of sites is selected across multiple replicates,and epidemic parameters. Across all *R*_0_ and replicates, the distribution is heavily concentrated at small number of selected sites, indicating that only a limited number of sites-typically fewer than 6, are sufficient to achieve near-optimal forecast performance. We also observe that, as epidemic intensity increases (*R*_0_ = 4), the distribution becomes wider, suggesting that scenarios with higher transmission levels, may benefit from a slightly larger subset of sites. Overall, the results highlight that short term robust hospitalization forecasting can be achieved with sparse, information-rich site selections rather than large surveillance networks.

**Figure 6.**
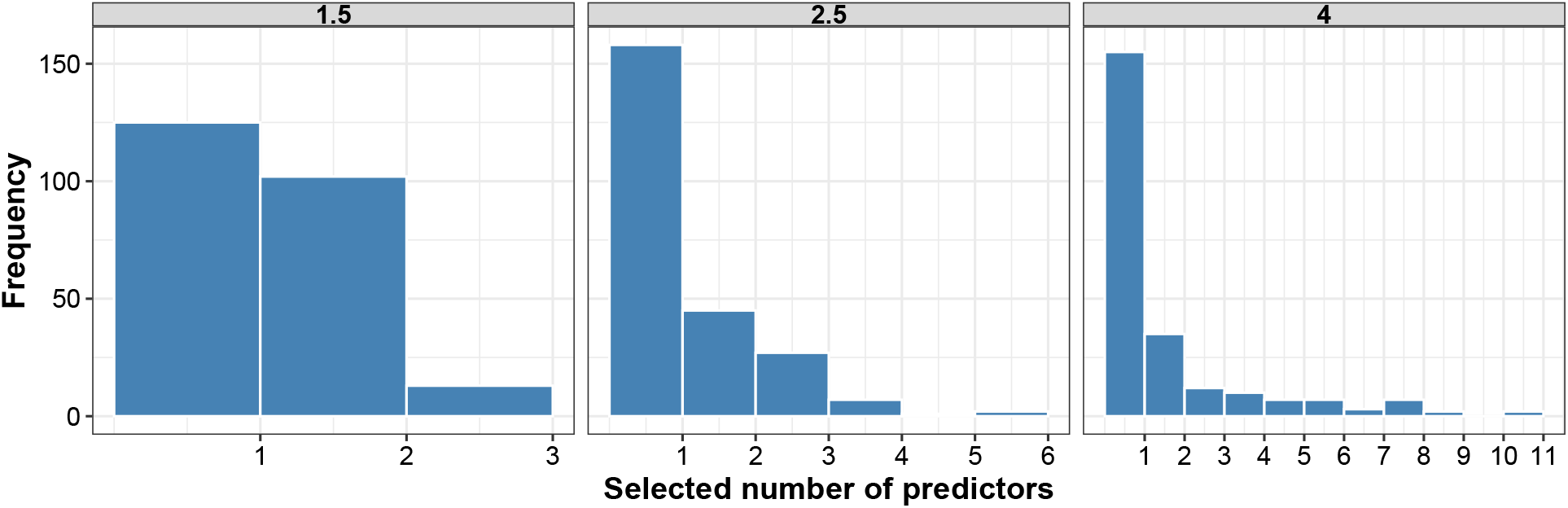
Frequency distribution of optimal number of sites across 60 replicates for *R*_0_ ∈ {1.5, 2.5, 4}.

#### 6.2 Rolling Window Forecast Evaluation and Model Complexity Comparison

We additionally compared our forecasting approaches to a standard LASSO regression model incorporating wastewater predictors. To assess and contrast model complexity across approaches, we quantified the number of predictors selected per rolling evaluation window. We focus on two multivariate strategies: (i) LASSO, which performs regularized variable selection independently at each rolling window, and (ii) a greedy ARIMA-X procedure, which sequentially adds predictors only when their inclusion yields an improvement in forecast accuracy relative to an autoregressive baseline.

Across replicates spanning different values of the basic reproduction number (*R*_0_) and replicates, the LASSO model consistently selected a large number of wastewater predictors. Averaged over rolling windows beginning at week 30, when both hospitalization outcomes and wastewater viral load measurements are jointly available – the LASSO model selected approximately 19 predictors per evaluation window, with the 90th percentile exceeding 31 predictors. This indicates that LASSO frequently distributes predictive weight across a broad set of correlated wastewater signals, resulting in relatively high-dimensional models. Most commonly selected sites with LASSO also have moderate positive correlations (ranging between 0.3 and 0.6) with hospitalization counts.

In contrast, the greedy ARIMA-X procedure selected very few predictors across replicates and across multiple *R*_0_ settings. Averaged over all replicated the model typically selected 1-10 sites. By construction, the greedy ARIMA-X approach retains a predictor only when its inclusion demonstrably improves out-of-sample forecast performance beyond that achieved by autoregressive dynamics alone. The fact that only a small number of predictors are consistently selected suggests that much of the wastewater signal is either redundant with the temporal autocorrelation already captured by the ARIMA component, or too weak and unstable to yield reliable improvements in short-term forecasts. Table 1 shows the average RMSE across all replicates from the three methods. LASSO and greedy ARIMA-X are generally better than ARIMA across all horizons. Figure 7 shows that the greedy ARIMA-X model consistently achieves lower or comparable error using a sparse subset of selected sites compared to the LASSO-based model. These results show that the greedy ARIMA-X outperforms baseline ARIMA model significantly during periods of rapid surge, while maintaining performance comparable to LASSO. These results indicate that the greedy approach effectively isolates the most informative wastewater signals during high-growth periods without incurring the increased model complexity associated with LASSO.

**Table 1:**
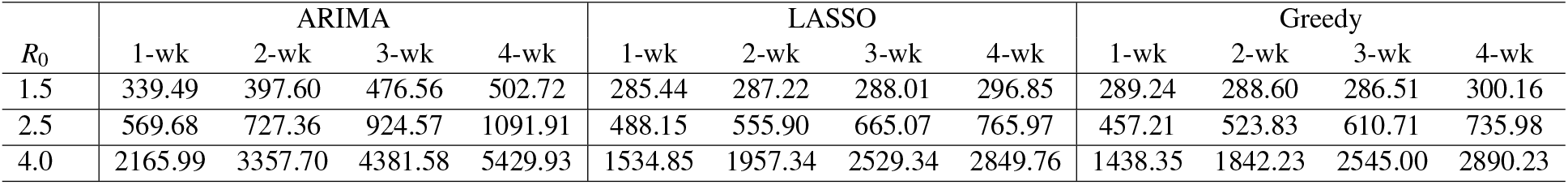
RMSE comparison of baseline ARIMA, LASSO, and Greedy (ARIMA-X) models across forecast horizons and *R*_0_ values.

| $R_0$ | ARIMA | | | | LASSO | | | | Greedy | | | |
| --- | --- | --- | --- | --- | --- | --- | --- | --- | --- | --- | --- | --- |
|  | 1-wk | 2-wk | 3-wk | 4-wk | 1-wk | 2-wk | 3-wk | 4-wk | 1-wk | 2-wk | 3-wk | 4-wk |
| 1.5 | 339.49 | 397.60 | 476.56 | 502.72 | 285.44 | 287.22 | 288.01 | 296.85 | 289.24 | 288.60 | 286.51 | 300.16 |
| 2.5 | 569.68 | 727.36 | 924.57 | 1091.91 | 488.15 | 555.90 | 665.07 | 765.97 | 457.21 | 523.83 | 610.71 | 735.98 |
| 4.0 | 2165.99 | 3357.70 | 4381.58 | 5429.93 | 1534.85 | 1957.34 | 2529.34 | 2849.76 | 1438.35 | 1842.23 | 2545.00 | 2890.23 |

**Figure 7.**
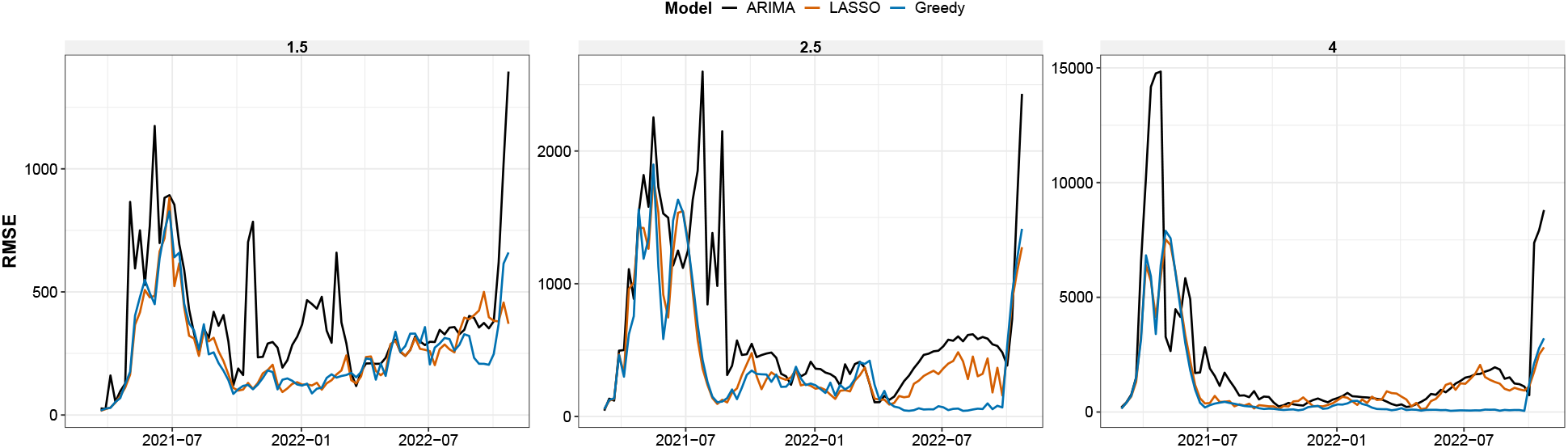
Temporal evolution of forecasting error (RMSE) for baseline ARIMA, LASSO, and greedy ARIMA-X models using Wastewater Digital Twin data. Lines show weekly out-of-sample RMSE across forecast horizons for each modeling approach, with facets corresponding to different epidemic scenarios.

## 7 DISCUSSION AND CONCLUSION

We present a systematic framework for optimizing wastewater surveillance site selection to support short-term hospitalization forecasting. Using both real-world surveillance data and a wastewater digital twin, we demonstrate that accurate forecasts can generally be achieved using a small, carefully selected subset of wastewater treatment plants, rather than broad population coverage. While wastewater viral activity levels are often moderately correlated with hospitalization dynamics, their marginal contribution to forecasting accuracy beyond autoregressive structure is limited. Our comparison with LASSO-based and ARIMA-X models highlight an important trade-off between modeling complexity and forecast relevance.

### Limitations

Firstly, both the greedy and LASSO models assume primarily linear relationships between wastewater signals and hospitalizations, and therefore may not fully capture nonlinear dependencies or spatial and neighborhood-level interactions among sites. Further, wastewater viral load measurements are subject to noise, reporting delays, and variability in sampling frequency and laboratory protocols across states and facilities, which may affect the stability and transferability of selected site subsets.

The simulation analysis with digital twin data presented here is limited to a finite set of epidemic scenarios characterized by three values of the basic reproduction number (*R*_0_). While these scenarios capture a range of transmission intensities, future work will extend this framework to explore a broader spectrum of epidemic conditions, including alternative parameter settings, multiple pathogen variants, and importation-driven outbreaks. Second, digital twin model relies on simplified representations of transmission, surveillance process and thus require future work incorporating more scenarios and uncertainty analysis with the wastewater data to with the site selection model.

## Data Availability

All data produced are available online at the links provided.

https://data.cdc.gov/Public-Health-Surveillance/CDC-Wastewater-Data-for-SARS-CoV-2/j9g8-acpt/about_data

https://data.cdc.gov/Public-Health-Surveillance/Weekly-United-States-COVID-19-Hospitalization-Metr/akn2-qxic/about_data}

## 8 ACKNOWLEDGEMENTS

The authors would like to thank the members of the Biocomplexity Institute for their valuable discussions and support. This work was partially supported by the Centers for Disease Control and Prevention (CDC) through the Pathogen Genomics Centers of Excellence (PGCoE) Network under Grant No. 6NU50CK000555-03-01, and by the National Science Foundation (NSF) through the Expeditions in Computing Program under Grant No. CCF-1918656.

## Footnotes

1 https://data.cdc.gov/Public-Health-Surveillance/NWSS-Public-SARS-CoV-2-Concentration-in-Wastewater

2 https://data.cdc.gov/Public-Health-Surveillance/Weekly-United-States-COVID-19-Hospitalization-Metr/akn2-qxic/about_data

